# Evidence of health-system resilience in the uptake of antiretroviral therapy in Sierra Leone during the COVID-19 pandemic: a nationwide retrospective study

**DOI:** 10.1101/2025.03.12.25323867

**Authors:** Jarieu Tucker, Joseph Marah, Ginika Egesimba, Zeleke Abebaw Mekonnen, Hannah Sheriff, Bailah Molleh, Stephen Sevalie, Adrienne K. Chan, Sharmistha Mishra, Sulaiman Lakoh

**Affiliations:** Ministry of Health, Government of Sierra Leone, Freetown, Sierra Leone; Research and Scientific Division, Sustainable Health Systems, Freetown, Sierra Leone; Research Unit, Partners in Health, Koidu, Sierra Leone; College of Medicine and Allied Health Sciences, University of Sierra Leone, Freetown, Sierra Leone; 34 Military Hospital, Republic of Sierra Leone Armed Forces, Wilberforce, Freetown, Sierra Leone; Division of Infectious Diseases, Department of Medicine, Faculty of Medicine, University of Toronto, Toronto, Canada

**Keywords:** COVID-19, HIV, antiretroviral therapy, ART, Sierra Leone, health-system strengthening, health system resiliency

## Abstract

**Background:** Sustained access to and uptake of antiretroviral therapy (ART) among people living with HIV is a critical component of the HIV response. Limited data exist evaluating the extent to which the COVID-19 pandemic may have disrupted HIV treatment services in Sierra Leone. This study aims to describe differential patterns in the uptake of ART before, during, and after the COVID-19 emergency period in the country.

**Methods:** We conducted a retrospective cross-sectional study using aggregated secondary program data extracted from the national health information system to reflect the uptake of ART in three time periods: March 2019-February 2020 (pre-COVID-19); March 2020-February 2021 (during COVID-19) and March 2021- February 2022 (post COVID-19). Outcomes were compared across the three time periods and stratified by sex and by region.

**Results:** There was a steady increase in the overall ART uptake from 365,326 pre-COVID-19 to 416,069 during COVID-19 and 518, 426 post-COVID-19, which masks the more significant relative decline affecting new ART initiations due to COVID-19: pre-COVID-19 (8, 958), during COVID-19 (8,777), and post-COVID-19 (13, 996). A sharp decline in new ART initiation was consistent with the surge in COVID-19, reflected geographical variability, but showed no sex difference.

**Conclusion:** Variations on the impact of COVID-19 across regions underscores the need for targeted interventions to mitigate the impact of future structural shocks on HIV services. The steady increase in overall uptake of ART suggests resiliency to a significant disruption and reflects pre-emptive health systems adaptations. The notable increase in new initiations in the post-COVID-19 period could suggest catch-up of delayed new initiations or more also reflect increasing HIV incidence and the critical importance of sustained investments for resiliency.

## Introduction

The HIV epidemic in Sierra Leone remains a persistent challenge, with variations in HIV prevalence across districts and population groups [1, 2]. HIV prevalence rates increased from 1.7% in 2013 to 1.73% in 2019, giving an estimated 77,000 people living with HIV in the country in 2023 [1–3].

Sustained antiretroviral therapy (ART) is critical to transforming the course of HIV infection from a life-threatening progressive disease to a chronic, manageable health condition and to reduce population-level HIV transmission [4]. To achieve this goal, UNAIDS has recommended the “95- 95-95” target, wherin, by the end of 2025, 95% of people living with HIV will know their infection status, 95% of those who know their status will receive ART, and 95%of those on treatment will achieve sustained viral suppression [5,6].

Despite this global recommendation, there are significant gaps in meeting the UNAIDS “95-95- 95” targets in Sierra Leone. Only 83% of people living with HIV in the country are aware of their diagnosis. Only 83% of people diagnosed with HIV receive treatment, and of these, only 65% achieve sustained viral suppression [3]. The consequence of the gaps in the cascade of HIV care in Sierra Leone is reflected in the high HIV prevalence of 24% in hospital settings, high prevalence of advanced HIV diseases amongst young people, and a high in-hospital mortality of 30.1% [7–9].

As efforts were being made to fill the gaps in the HIV cascade, the COVID-19 pandemic hit and challenged the gains in the health system in Sierra Leone and around the world [10–14]. Sierra Leone reported its first case of COVID-19 on March 30, 2020, although restrictive public health measures had already been in place in January 2020 [11]. Subsequently, the Government of Sierra Leone instituted measures to reduce the transmission of SARS-CoV-2, such as lockdowns, movement restrictions, and redirecting the health infrastructure and workforce to combat COVID-19. While these measures were aimed at controlling the spread of COVID-19, they also disrupted the delivery of essential services [11–14]. To mitigate the impact of the COVID-19 pandemic, the National AIDS Control Program implemented policies to ensure the continued provision of treatment services. These include decentralized medication delivery, multi-month (rather than monthly) prescribing and dispensing of HIV medications, and follow-up phone calls (rather than face-to-face care) to track adherence, clinical care, and treatment refills. To date, there is no published literature on the impact of the COVID-19 pandemic on HIV treatment uptake in Sierra Leone.

This study aims to assess patterns of ART uptake before, during, and after the COVID-19 emergency period in Sierra Leone to understand the resiliency of its health system with these adaptations, inform future pandemic preparedness, and support a sustainable HIV response.

## Methods

### Study design and population

We conducted a retrospective cross-sectional study of all persons aged 15 years or older diagnosed with HIV who were receiving ART between February 2019 and February 2022 using aggregated secondary program data. New ART uptake was represented by people living with HIV who were newly diagnosed and started ART during the reporting month. Overall ART treatment uptake refers to both treatment uptake by newly initiated ART and treatment uptake by people with HIV who have previously received ART.

### Study setting

Sierra Leone has a population of 7.5 million and is comprised of 16 districts administered under five regions: Eastern (Kailahun, Kenema, and Kono districts), Western Area (Urban and Rural districts), Northern (Bombali, Falaba, Koinadugu and Tonkolili districts), Northwestern (Kambia, Karene, and Port Loko districts) and Southern (Bo, Bonthe, Moyamba, Pujehun districts). There are large disparities between districts in the number of people living with HIV receiving treatment [15].

The National AIDS Control Program leads the health sector response to HIV. It coordinates the response across the five regions of Sierra Leone. HIV treatment services are provided in 826 primary, secondary and tertiary health facilities in Sierra Leone. Health providers operate HIV clinics on daily basis, offering refill services, including treatment readiness and new treatment initiations. Antiretroviral drugs are dispensed using multi-month scripting with clients established on ART receiving three to six months supplies. New clients on ART or those not established on ART receive monthly supply of medicines.

### Data sources and variables

Data was extracted in August 20, 2023 from the District Health Management System 2 (DHIS2), a national health management information systems database consisting of secondary aggregate data collected monthly from health facilities providing HIV services. Information collected from patients is aggregated and transmitted to the districts in a de-identified format and then entered in the DHIS2. Data recording and reporting primarily use a paper-based system, except for a few facilities that have hybrid systems. The data collection tools in the DHIS2 have been standardized across the country, and the information collected is aligned with national indicators. As part of the routine DHIS2 data management system, data verification was carried out at the health facility and districts before data was entered into the DHIS2, and data checks are performed quarterly to improve data quality. As part of this study, we extracted 36 months of aggregated data on ART uptake, disaggregated by sex and region, to answer the study objectives. There are no missing data for any month.

We extracted monthly data from March 2019 to February 2022 to reflect the uptake of ART in three time periods: March 2019-February 2020 (pre-COVID-19, 12 months); March 2020- February 2021(during COVID-19, 12 months); March 2021- February 2022 (post-COVID-19, 12 months). Variables extracted included sex, district, region, and the outcome of interest: total number of people living with HIV in the reporting month and number people newly diagnosed with HIV initiated on treatment in the reporting month.

### Data management and analysis

Data in Excel format was cleaned and coded prior to analysis. We described monthly trends in the absolute number of ART uptake (new and overall) using Excel to describe national and regional variation before, during, and post-COVID-19 pandemic, and stratified by sex and region. Descriptive statistics were used to summarize the results.

### Ethics

Ethical approval was obtained from the Sierra Leone Ethics and Scientific Review Committee (approval number 021/05/2023). As the study involved secondary analysis of routine, anonymized, and aggregated program data, informed consent was waived, and the research team did not have access to information that could identify individual participants during or after data collection.

## Results

### Trends in the uptake of ART before, during and after COVID-19

There was a steady increase in the overall ART uptake from 365,326 pre-COVID-19 to 416,069 during COVID-19 and 518, 426 post-COVID-19, which masks the more significant relative decline affecting new ART initiations due to COVID-19: pre-COVID-19 (8, 958), during COVID-19 (8,777), and post-COVID-19 (13, 996) (Table 1). Except for a transient decline between June 2021 and July 2021, there was a steady increase in the overall uptake of ART across the three time periods, maintaining an upward trend pre, during and post-COVID-19 pandemic (Figure 1). The number of new ART initiations also increased over time (Table 1, Figure 2), but there was a sharp decline during the early period of COVID-19 (March-April 2020) with a rapid recovery and ongoing increases in new ART initiations in the post-COVID-19 period (Figure 2). The largest proportion of new ART initiations were among women, and in the Western Area (Table 1), and the pattern of the temporary decline and recovery in new ART initiations were consistent across sex (Figure 2) and regions (Figure 3). The Northwest saw increases in new ART initiations in the post-COVID-19 period, departing from earlier trends similar to other regions outside the Western Area.

**Figure 1.**
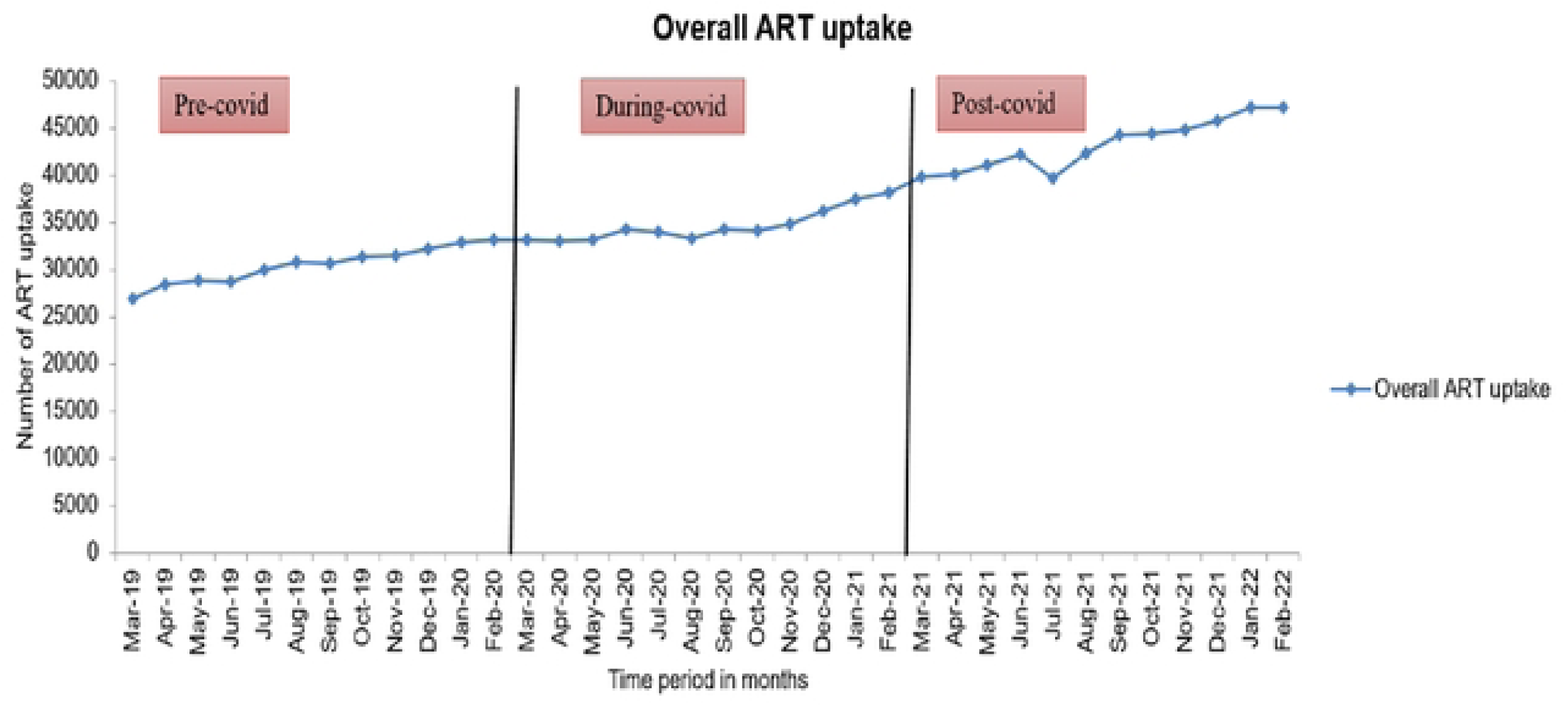
Uptake of ART before, during and after the COVID-19 pandemic. Pre-COVID- (March 2019-Feb 2020), during COVID- (March 2020-Feb 2021), Post COVID-(March 2021-Feb 2022) ART-Antiretroviral Therapy

**Figure 2.**
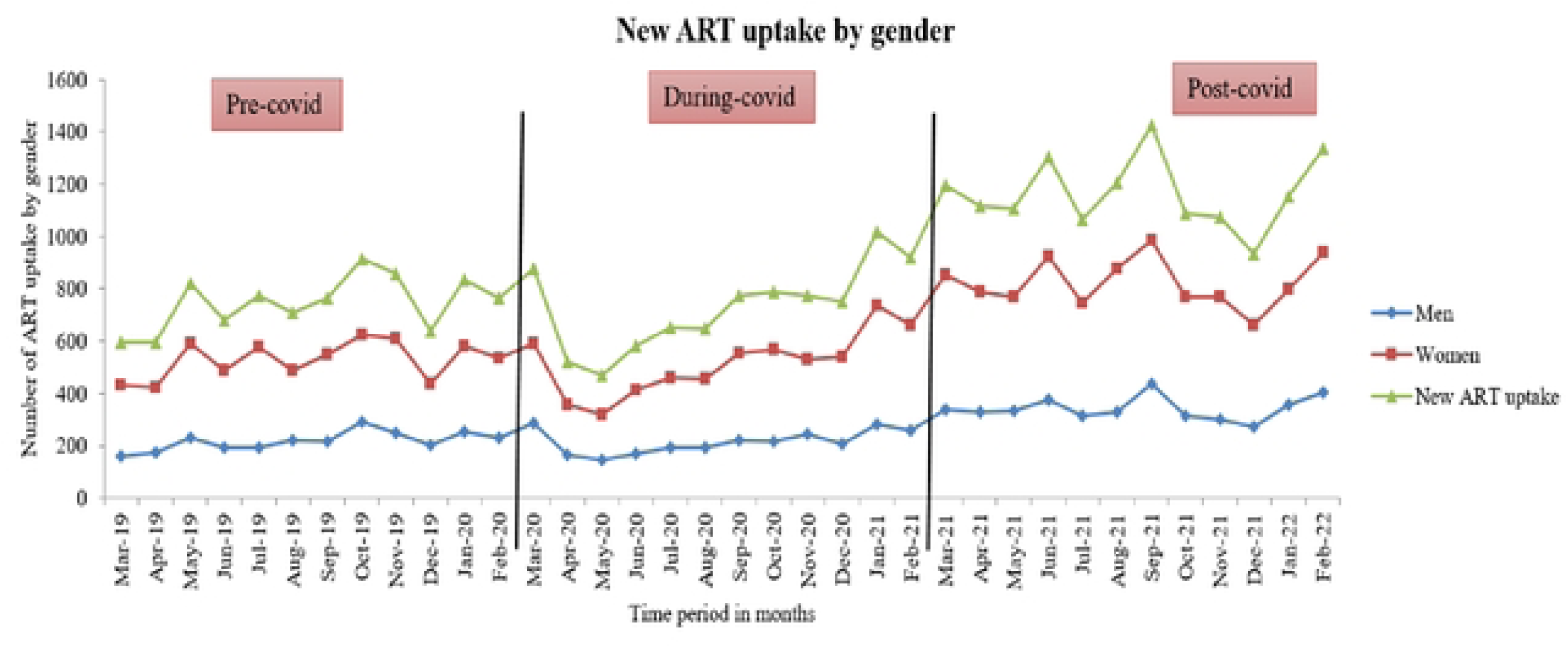
Uptake of ART among newly diagnosed PLHIV in Sierra Leone by gender March 2019-Febuary 2022. Pre-COVID- (March 2019-Feb 2020), during COVID- (March 2020-Feb 2021), Post COVID-(March 2021-Feb 2022) ART-Antiretroviral Therapy

**Figure 3:**
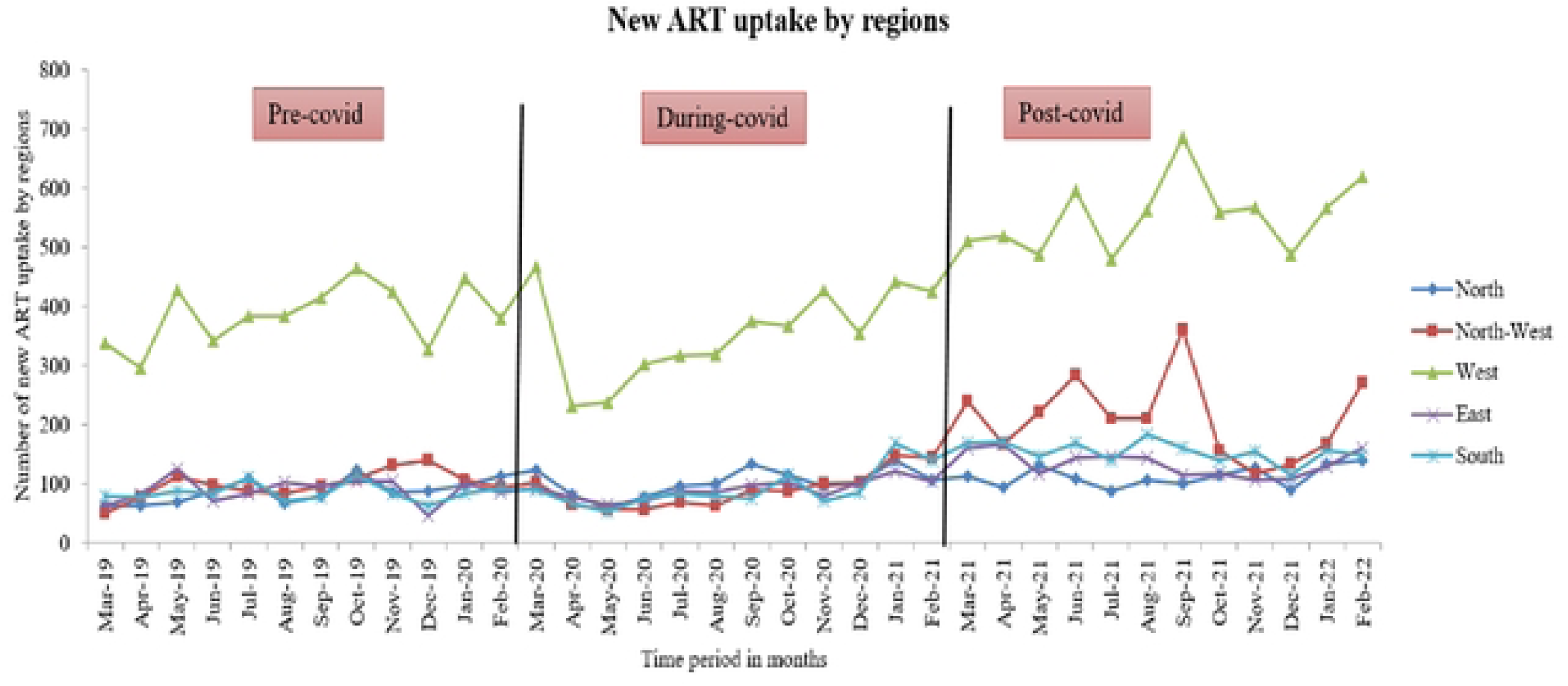
ART uptake among newly diagnosed PLHIV within the five regions in Sierra Leone. Pre-COVID- (March 2019-Feb 2020), during COVID-(March 2020-Feb 2021), Post COVID-(March 2021-Feb 2022) ART-Antiretroviral Therapy

**Table 1:** Uptake of ART pre, during and post COVID-19 in Sierra Leone.

| Data description | Time period |  |  |
| --- | --- | --- | --- |
|  | Pre-COVID-19<br>March 2019-February 2020,<br>12 months | During-COVID-19<br>March 2020-February<br>2021, 12 months | Post-COVID-19<br>March 2021-February 2022,<br>12 months |
| **Total number who received ART during the 12 month period** |  |  |  |
| **Previously on ART** | 356368 | 407292 | 504430 |
| **Newly initiated on ART** | 8958 | 8777 | 13996 |
| **Overall** | 365326 | 416069 | 518426 |
| **Number newly initiated on ART by gender, during the 12 month period** |  |  |  |
| **Men** | 2621 | 2590 | 4118 |
| **Women** | 6337 | 6187 | 9878 |
| **Total** | 8958 | 8777 | 13996 |
| **Number newly initiated on by region, during the 12 month period** |  |  |  |
| **North** | 1048 | 1234 | 1348 |
| **North-West** | 1189 | 1087 | 2539 |
| **Western** | 4629 | 4260 | 6637 |
| **East** | 1066 | 1090 | 1618 |
| **South** | 1026 | 1106 | 1854 |
| **Total** | 8958 | 8777 | 13996 |

Pre-COVID- (March 2019-Feb 2020), during COVID- (March 2020-Feb 2021), Post COVID-(March 2021-Feb 2022) ART-Antiretroviral Therapy

## Discussion

This study assessed the impact of COVID-19 on ART uptake in all five regions of Sierra Leone and highlights five key findings. First, there was a sharp decline in the new uptake of ART among people living with HIV who were newly diagnosed in all five regions of Sierra Leone at the start of COVID-19 in March 2020. Second, the decline in the uptake of new ART services was followed by a gradual recovery and steady increase of new ART uptake during COVID-19 and continued despite intermittent disruptions in services coinciding with the Delta wave at the end of 2020 and the Omicron wave June and July 2021. Third, most of the changes were driven by uptake patterns in the Western Area, which includes the capital, Freetown, and where a significant number of people living with HIV access services. Furthermore, the sharpest decline and nadir in ART uptake occurred in April 2020, after the first case of COVID-19, coinciding specifically with inter-country travel restrictions and lockdowns. Finally, except for a transient decline in June 2021 and July 2021, there was a steady increase in the overall uptake of ART across the three time periods, maintaining an upward trend before and during and after the COVID-19 pandemic.

There is a higher impact of COVID-19 on the uptake of services in people living with HIV newly initiated on ART than those already on treatment. These changes are likely reflective of planned programmatic mitigations to ensure those already on treatment remained in care with multi-month supply in anticipation of restrictions, the use of mobile phones for remote monitoring, and a greater client awareness of the critical importance of ensuring and anticipating uninterrupted supply. The decline in the uptake new ART is also reflective of a preceding nadir in HIV testing services that occurred in January 2020, which acts as the gateway to the uptake of new ART [13].

Although this is the first study in Sierra Leone to report the impact of COVID-19 on the national uptake of ART services, it reflects the global pattern of disruptions in essential services by the COVID-19 pandemic related to lockdowns at the beginning of the pandemic and underscores the need to protect essential health services such as HIV during public health emergencies [16, 17].

In Sierra Leone, decentralized drug delivery, multi-months scripting and dispensing of HIV medicines and follow up calls to track adherence, clinical care, and treatment refills were adapted by the national program to mitigate the impact of the pandemic. Our study indicated intermittent declines in new ART uptake coincided with peaks of COVID 19 pandemics probably reflecting supply chain problems, movement restrictions, closure of health facilities and phobia of patients to visit health clinics.

The notable resilience in the ART program during and after COVID-19 may reflect on these mitigation strategies but there is limited evidence in the literature to compare the resilience of our treatment program, as most studies are restricted to the pandemic period. The IeDEA West Africa Cohort reported variable findings on the impact of COVID-19 on the uptake of new ART. Clinics in Burkina Faso and Côte d’Ivoire had no change in the number of new ART initiations during the pandemic, but the Nigerian IeDEA cohort of new initiates of ART decreased during the pandemic [18]. A time series analysis in Botswana on national HIV data comparing April 2019-March 2020 (pre-COVID-19) to March 2020-April 2021 (during COVID-19) reported a significant impact of the COVID-19 with a 43% reduction in ART initiations [19]. A study in South Africa reported a decline of 46% in weekly ART initiations in March 2020-June 2020 (during lockdown) [20]. Similar reductions in the uptake of ART were also reported in Tanzania, Kenya, Uganda and Nigeria [21]. Dorward and Ben Farhat et al. reported recovery of ART collections following easing of the lockdown during the COVID-19 pandemic [18, 20].

There are geographical differences in the uptake of new ART, requiring attention to national mitigation planning. Again, the result of this study demonstrates that the challenges of accessing ART are significantly higher in men compared to women. However, unlike other studies in Africa, in our study, the impact of COVID-19 on ART did not differ significantly between sex [22].

Reflecting on these gaps in the current geopolitical context highlights some concerns and the critical need for a rapid evaluation of national programmatic outcomes in response to disruptions from the current HIV funding freeze by the United States Government [23]. The increase in the overall ART uptake in the post-COVID-19 period in our study may reflect a catch up strategy to mitigate the impact of the COVID-19 pandemic or perhaps it may be driven by the increasing incidence of HIV. The resiliency in HIV service delivery demonstrated in our study is likely contingent on stable and long-term investments particularly by the US government which had contributed 85.1% of the global HIV overseas development aid to Sierra Leone, putting it as the top 7th country recipient of US global HIV funding as a share of the total Global HIV Funding, 2021-23 [24]. The HIV uptake of ART services in the post-COVID-19 period in our study raises some concerns about sustainability in the current context. We call on the Government of Sierra Leone to expedite the development and implementation of an HIV sustainability road map.

Our study has strengths and limitations. The use of data from all public and private facilities providing HIV services in Sierra Leone reflects a national representation of the HIV treatment program. Our use of long-term routine data over a large period considers trends across the period in the use of ART and allows more accurate quantification of the effect over the total period of the pandemic. This study contributes to the broader understanding of pandemic preparedness in chronic disease management and advocates for adaptive healthcare policies that prioritize equitable access to essential services during health emergencies.

We used secondary data in DHIS2 and have no control over the quality of the data input into this database. Furthermore, the use of aggregate data limited our ability to analyse relevant variables such as age groups and sub-populations.

## Conclusion

We report that when the COVID-19 outbreak began in March 2020, there was a sharp drop in the number of people receiving new ART in all five regions of Sierra Leone. The decline in the uptake of new ART was followed by a gradual recovery, even during the pandemic, and this upward trend continued despite intermittent declines during the Delta wave in late 2020 and the Omicron wave in June 2021 and July 2021. Most of the changes in the uptake of new ART are driven by patterns of uptake in Western Area, but did not show clear gender differences. These findings provide critical insights into the challenges faced but also addressed by Sierra Leone’s healthcare system in maintaining HIV treatment services amidst the COVID-19 crisis. By highlighting variations across regions and demographic groups, the study underscores the need for targeted interventions to mitigate the impact of future pandemic impact on HIV services but also highlights the critical need for sustained investments to ensure programmatic resiliency to unexpected disruptions.

## Data Availability

The datasets used and/or analyzed for this study are available from https://figshare.com/account/items/28574516/edit.

## Declaration section

The authors declare no conflict of interest

## Acknowledgements

This research was conducted through a partnership between Sustainable Health Systems Sierra Leone, and the Li Ka Shing Research Institute, Unity Health, University of Toronto, Canada. The training model used that resulted in this publication was adapted from the Structured Operational Research and Training Initiative (SORT IT), a global partnership led by the Special Programme for Research and Training in Tropical Diseases at the World Health Organization (WHO/TDR, Geneva, Switzerland), for which SL, AKC and SM are SORT-IT Mentors. Mentorship was provided by Sustainable Health Systems, (Freetown, Sierra Leone); Ministry of Health, Government of Sierra Leone; University of Toronto (Toronto, Canada); Partners in Health Sierra Leone (Koidu, Sierra Leone). The authors would like to acknowledge Kristy Cheuk Yin Yiu and Bailah Molleh for Project Management and Coordination.

## Funding

This research was funded by the Canadian Institutes for Health Research (CIHR) through an Operating Grant (WI1-179883) Addressing the Health Impacts of COVID-19. SM is funded by a Tier 2 Canada Research Chair in Mathematical Modeling and Program Science (CRC File Number 950-232643). The funders had no role in the study design, data collection and analysis, decision to publish, or preparation of the manuscript.

## Conflict of interest

None to declare

## Author’s contribution

Conceptualization and study design: JT, HS, GE and SL

Data curation and validation: JT, HS, GE, BM, JM, SM

Methodology and formal analysis: JT, HS, GE, JM, ZA, BM, SM, Funding acquisition, resources, supervision: SS, SL, SM, AKC

Writing – original draft preparation: JT, HS, GE, JM, ZA, BM, AKC and SM

Writing – review & editing: SL, SS, AKC and SM

## References

1. SLDHS. (2019). Sierra Leone Demographic and Health Survey 2019. Freetown, Sierra Leone, and Rockville, Maryland, USA: Stats SL and ICF. Accessed on: https://www.statistics.sl/images/StatisticsSL/Documents/DHS2018/sldhs2019kir.pdf.

2. Sierra Leone Demographic Health Survey (SLDHS). 2019. Available at: https://dhsprogram.com/pubs/pdf/PR122/PR122.pdf. (Accessed on February 10 2025)

3. UNAIDS Facts sheet (2023)-Available at: https://www.unaids.org/en/regionscountries/countries/sierraleone. Accessed September 20, 2024.

4. Jelliman P, Porcellato L. HIV is Now a Manageable Long-Term Condition, But What Makes it Unique? A Qualitative Study Exploring Views About Distinguishing Features from Multi-Professional HIV Specialists in Northwest England. J Assoc Nurses AIDS Care. 2017 Jan-Feb;28(1):165–178. doi: 10.1016/j.jana.2016.09.008. Epub 2016 Sep 22. PMID: 27836199.

5. Frescura L, Godfrey-Faussett P, Feizzadeh A A, El-Sadr W, Syarif O, Ghys PD; on and behalf of the 2025 testing treatment target Working Group. Achieving the 95 95 95 targets for all: A pathway to ending AIDS. PLoS One. 2022 Aug 4;17(8): e0272405. doi: 10.1371/journal.pone.0272405. PMID: 35925943; PMCID: PMC9352102.

6. UNAIDS. Understanding Fast-Track: accelerating action to end the AIDS epidemic by 2030. 2015. Available from: unaids.org/sites/default/files/media_asset/201506_JC2743_Understanding_FastTrack_en.pdf.

7. Lakoh S, Firima E, Jiba DF, Sesay M, Conteh MM, Deen GF. Low partner testing in high HIV prevalence setting in Freetown, Sierra Leone: a retrospective study. BMC Res Notes. 2019 Sep 24;12(1):629. doi: 10.1186/s13104-019-4662-9. PMID: 31551091; PMCID: PMC6760048.

8. Baldeh M, Kizito S, Lakoh S, Sesay D, Williams SA, Barrie U, Dennis F, Robinson DR, Lamontagne F, Amahowe F, Turay P, Bahar OS, Geng E, Ssewamala FM. Advanced HIV disease and associated factors among young people aged 15-24 years at a tertiary hospital in Sierra Leone: a cross-sectional study. BMC Infect Dis. 2024 June 20;24(1):611. doi: 10.1186/s12879-024-09524-5. PMID: 38902606; PMCID: PMC11191260.

9. Lakoh S, Jiba DF, Kanu JE, Poveda E, Salgado-Barreira A, Sahr F, Sesay M, Deen GF, Sesay T, Gashau W, Salata RA, Yendewa GA. Causes of hospitalization and predictors of HIV-associated mortality at the main referral hospital in Sierra Leone: a prospective study. BMC Public Health. 2019 Oct 21;19(1):1320. doi: 10.1186/s12889-019-7614-3. PMID: 31638941; PMCID: PMC6805411.

10. Prabhu S, Poongulali S, Kumarasamy N. Impact of COVID-19 on people living with HIV: A review. J Virus Erad. 2020 Nov;6(4):100019. doi: 10.1016/j.jve.2020.100019. Epub 2020 Oct 15. PMID: 33083001; PMCID: PMC7560116.

11. Lakoh S, Jiba DF, Baldeh M, Adekanmbi O, Barrie U, Seisay AL, Deen GF, Salata RA, Yendewa GA. Impact of COVID-19 on Tuberculosis Case Detection and Treatment Outcomes in Sierra Leone. Trop Med Infect Dis. 2021 Aug 19;6(3):154. doi: 10.3390/tropicalmed6030154. PMID: 34449755; PMCID: PMC8396336.

12. MacIntyre CR, Heslop DJ. Public health, health systems and palliation planning for COVID-19 on an exponential timeline. Med J Aust. 2020;212(10):440–442.e1. doi: 10.5694/mja2.50592

13. Lakoh S, Bangura MM, Adekanmbi O, Barrie U, Jiba DF, Kamara MN, Sesay D, Jalloh AT, Deen GF, Russell JBW, Egesimba G, Yendewa GA, Firima E. Impact of COVID-19 on the Utilization of HIV Testing and Linkage Services in Sierra Leone: Experience from Three Public Health Facilities in Freetown. AIDS Behav. 2024 Apr;28(4):1235–1243. doi: 10.1007/s10461-023-04149-2. Epub 2023 Aug 29. PMID: 37642824; PMCID: PMC10940454.

14. Sevalie S, Youkee D, van Duinen AJ, Bailey E, Bangura T, Mangipudi S, Mansaray E, Odland ML, Parmar D, Samura S, van Delft D, Wurie H, Davies JI, Bolkan HA, Leather AJM. The impact of the COVID-19 pandemic on hospital utilisation in Sierra Leone. BMJ Glob Health. 2021 Oct;6(10): e005988. doi: 10.1136/bmj-2021-005988. PMID: 34635552; PMCID: PMC8506048.

15. Statistics Sierra Leone-2021 Mid-Term Population and Housing Census. Available at: https://www.statistics.sl/images/StatisticsSL/Documents/Census/MTPHC_Preliminary_Report/Final_Preliminary_Report_2021_MTPHC.pdf. Accessed September 27, 2024.

16. Adepoju P. Tuberculosis and HIV responses threatened by COVID-19. Lancet HIV. 2020;7: e319–e320. doi: 10.1016/S2352-3018(20)30109-0.

17. Nachega JB, Kapata N, Sam-Agudu NA, Decloedt EH, Katoto PDMC, Nagu T, Mwaba P, Yeboah-Manu D, Chanda-Kapata P, Ntoumi F, Geng EH, Zumla A. Minimizing the impact of the triple burden of COVID-19, tuberculosis and HIV on health services in sub-Saharan Africa. Int J Infect Dis. 2021 Dec;113 Suppl 1: S16-S21. doi: 10.1016/j.ijid.2021.03.038. Epub 2021 Mar 20. PMID: 33757874; PMCID: PMC7980520.

18. Ben Farhat J, Tiendrebeogo T, Malateste K, Poda A, Minga A, Messou E, Chenal H, Ezechi O, Ofotokun I, Ekouevi DK, Bonnet F, Barger D, Jaquet A; IeDEA West Africa Collaboration. Effects of the COVID-19 Pandemic on ART Initiation and Access to HIV Viral Load Monitoring in Adults Living with HIV in West Africa: A Regression Discontinuity Analysis. J Acquir Immune Defic Syndr. 2024 June 1;96(2):114–120. doi: 10.1097/QAI.0000000000003404. PMID: 38427928; PMCID: PMC11108739.

19. Sehurutshe A, Farooqui H, Chivese T. The Impact of COVID-19 on the HIV Cascade of Care in Botswana - An Interrupted Time Series. AIDS Behav. 2024 Aug;28(8):2630–2638. doi: 10.1007/s10461-024-04388-x. Epub 2024 Jun 10. PMID: 38856847; PMCID: PMC11286653.

20. Dorward J, KhuboneT, Gate K, Ngobese H, Sookrajh Y, Mkhize S, et al. The impact of the COVID-19 lockdown on HIV care in 65 South African primary care clinics: an interrupted time series analysis. Lancet HIV. 2021;8(3): e158–65. Epub 20210204.

21. Dear N, Duff E, Esber A, Parikh A, Iroezindu M, Bahemana E, et al. Transient reductions in human immunodeficiency virus (HIV) Clinic Attendance and Food Security during the Coronavirus Disease 2019 (COVID-19) pandemic for people living with HIV in 4 African countries. Clin Infect Dis. 2021;73(10):1901–5.

22. Bernard C, Hassan SA, Humphrey J, Thorne J, Maina M, Jakait B, Brown E, Yongo N, Kerich C, Changwony S, Qian SRW, Scallon AJ, Komanapalli SA, Enane LA, Oyaro P, Abuogi LL, Wools-Kaloustian K, Patel RC. Impacts of the COVID-19 pandemic on access to HIV and reproductive health care among women living with HIV (WLHIV) in Western Kenya: A mixed methods analysis. Front Glob Womens Health. 2022 Dec 12; 3:943641. doi: 10.3389/fgwh.2022.943641. PMID: 36578364; PMCID: PMC9790904.

23. Tram KH, Ratevosian J, Beyrer C. By executive order: The likely deadly consequences associated with a 90-day pause in PEPFAR funding. J Int AIDS Soc. 2025 Mar;28(3):e26431. doi: 10.1002/jia2.26431. PMID: 39996580; PMCID: PMC11851316.

24. Kaiser Family Foundation. The Trump Administration’s Foreign Aid Freeze and Global Health: The Biggest Gaps Left on the Donor Landscape. Available at: https://www.kff.org/global-health-policy/issue-brief/the-trump-administrations-foreign-aid-freeze-and-global-health-the-biggest-gaps-left-on-the-donor-landscape/. Accessed on March 8, 2025

